# Associations of plasma biomarkers of Alzheimer’s pathology with longevity and healthspan

**DOI:** 10.64898/2026.03.10.26348059

**Authors:** Bowei Zhang, Andrea Z. LaCroix, Susan M. Resnick, Steve Nguyen, Luigi Ferrucci, Towia A. Libermann, Long Ngo, Ramon Casanova, Alexander P. Reiner, Danni Li, Caroline M. Nievergelt, Adam X. Maihofer, Linda K. McEvoy, Aladdin H. Shadyab

**Affiliations:** Herbert Wertheim School of Public Health and Human Longevity Science, University of California San Diego, La Jolla, CA, USA; Laboratory of Behavioral Neuroscience, National Institute on Aging, National Institutes of Health, Baltimore, Maryland, USA; Department of Radiology, University of Pennsylvania, Philadelphia, PA, USA; Longitudinal Studies Section, Translational Gerontology Branch, National Institute on Aging, Baltimore, MD, USA; Division of Interdisciplinary Medicine and Biotechnology, Beth Israel Deaconess Medical Center, Boston, MA, USA; Beth Israel Deaconess Medical Center Genomics, Proteomics, Bioinformatics, and Systems Biology Center, Boston, MA, USA; Division of General Medicine, Department of Medicine, Beth Israel Deaconess Medical Center, Boston, MA, USA; Department of Biostatistics, Harvard T.H. Chan School of Public Health, Boston, MA, USA; Department of Biostatistics and Data Science, Wake University School of Medicine, Winston-Salem, NC, USA; Department of Epidemiology, University of Washington, Seattle, WA, USA; Department of Laboratory Medicine and Pathology, University of Minnesota, Minneapolis, MN, USA; Department of Psychiatry, University of California San Diego, La Jolla, CA, USA; Kaiser Permanente Washington Health Research Institute, Seattle, WA, USA; Division of Geriatrics, Gerontology, and Palliative Care, Department of Medicine, University of California San Diego, La Jolla, CA, USA

## Abstract

Plasma biomarkers of Alzheimer’s pathology have been studied in relation to cognitive decline and dementia, but no prior study has examined their associations with longevity or healthspan. In this cohort study of older women (N=2,576), we examined plasma biomarkers measured at baseline when women were cognitively unimpaired. We found that plasma biomarkers were associated with both exceptional longevity and healthspan, defined as survival to age 90 without cognitive impairment. Elevated levels of plasma p-tau217 (OR, 0.58; 95% CI, 0.50-0.68), p-tau181 (OR, 0.81; 95% CI, 0.71 – 0.93), NfL (OR, 0.70; 95% CI, 0.60 – 0.82) and GFAP (OR, 0.78; 95% CI, 0.68 – 0.90) were all associated with reduced odds of healthspan; findings survived adjustment for multiple comparisons. These findings suggest that plasma biomarkers may not only reflect ADRD pathology but also systemic aging processes that underlie lifespan and healthspan, underscoring their potential value as novel biomarkers of aging.

## INTRODUCTION

Life expectancy in the United States has significantly increased since the second half of the 20^th^ century. The estimated probability of surviving to age 90 or older increased from 6.1% for the 1949-51 birth cohort to 18.8% for the 2000 birth cohort.^1^ In 2020, individuals aged 90 and older comprised 3.4% of the population aged 65 and above, which is projected to reach 10% by 2050.^2,3^ Simultaneously, the aging of the population has been accompanied by an increase in the prevalence of Alzheimer’s disease (AD). As of 2025, an estimated 7.2 million Americans aged 65 and older are living with AD, which is projected to increase to 12.7 million by 2050.^4^ These trends underscore the growing need to identify determinants of healthspan - defined as the period of life spent in good health, free from major chronic diseases, functional limitations, and cognitive impairment - as well as novel biomarkers that track healthspan.

Plasma biomarkers have shown promise for detecting AD and related dementias (ADRD) in the preclinical stage.^5–9^ These include biomarkers of AD-specific pathology - such as phosphorylated tau at threonine 217 (p-tau217) and at threonine 181 (p-tau181) - ^6,7,10^ and non-specific markers of AD that signal broader neurological damage, including markers of neuronal injury (neurofilament light chain protein [NfL]) and of neuroinflammation (glial fibrillary acidic protein [GFAP]).^8–13^

Among cognitively healthy older adults, higher levels of pathology as indicated by these biomarkers are associated with cognitive decline and increased risk of dementia. ^8,10,12,14–17^ However, less is known about their relationship to cognitive health in late life (e.g., cognitively healthy longevity), an endpoint of relevance to the aging population.^18^ The distinction between longevity and cognitively healthy longevity is particularly important, because cognitive impairment is a primary cause of loss of independence in older adults, a feared outcome that is associated with high cost for affected individuals, their families, and society. There are currently no blood-based biomarkers capable of predicting who will survive to advanced age with intact cognition.

In this study, we examined associations of plasma ADRD biomarkers including p-tau217, p-tau181, NfL, and GFAP with (1) exceptional longevity (survival to age 90 years versus dementia-free death prior to age 90 years) and (2) cognitively healthy longevity (survival to age 90 years without mild cognitive impairment (MCI) or dementia versus dementia-free death prior to age 90 years), among participants from the Women’s Health Initiative Memory Study (WHIMS).

## METHODS

### Study design and population

WHIMS was designed to investigate the effects of hormone therapy on adjudicated cognitive outcomes (no impairment, mild cognitive impairment, or probable dementia) among 7,479 women ages 65-79 years who were cognitively unimpaired at randomization in 1996-1999. The WHIMS study design and detailed protocols are published elsewhere.^19–21^ Women with an intact uterus were randomly assigned to either conjugated equine estrogens (CEE) plus medroxyprogesterone acetate (MPA) vs placebo. Women with prior hysterectomy were randomly assigned to CEE alone vs placebo. The trials were stopped in 2002 and 2004, respectively, but annual in-person assessments for cognitive outcomes continued through 2007. In 2008, WHIMS transitioned to the WHIMS Epidemiology of Cognitive Health Outcomes (WHIMS-ECHO) study, which conducted annual telephone-administered to follow participants for the same cognitive outcomes through 2021.^22^

We examined a subset of 2,836 WHIMS women who were selected from WHIMS for assessment of baseline plasma ADRD biomarkers (see Supplementary Methods). Our analyses were restricted to women who had potential, because of birth year, to survive to age 90 at the end of the most recent WHI follow-up date of February 17^th^, 2024 (i.e., women born on or before February 17, 1934), and we also excluded women who died from dementia before reaching age 90 to rule out dementia-related death as a cause of our findings. Finally, only women with complete plasma biomarker information were included, resulting in a final analytic sample of 2,576 women (**Figure S1**).

The present study was approved by the Institutional Review Board of Fred Hutchinson Cancer Center. All women provided written informed consent.

### Plasma ADRD Biomarkers

Fasting blood was drawn at enrollment. Samples were processed, frozen at −70°C, and then shipped to a repository in Rockville, MD maintained by Fisher Bioservices. All plasma biomarker assays were performed at the Advanced Research and Diagnostic Laboratory (ARDL) at the University of Minnesota, USA, in 2024. Samples were shipped to the laboratory on dry ice. All blood samples were shipped to the lab in one batch to minimize batch effects. Levels of plasma biomarkers were measured using the Quanterix HD-X platform. The Simoa® Human Neurology 4-Plex E (N4PE) platform was used to measure levels of NfL and GFAP. P-tau181 was measured using the Simoa® p-tau181 v2 assay. P-tau217 was measured using the ALZpath Simoa® pTau-217 v2 assay. Samples were assayed in singlets with the inclusion of 192 duplicates; laboratory personnel were blinded to the inclusion of these duplicate samples and to cognitive impairment status. The average intra-assay coefficients of variation for NfL, GFAP, p-tau181, and p-tau217 derived from these duplicates were 7.1%, 9.8%, 10.2%, and 11.4%, respectively (Supplementary Methods).

### Longevity Outcomes

We first examined exceptional longevity, defined as survival to age 90, versus death before age 90 years. Death events were evaluated from baseline to February 17^th^, 2024, and were confirmed by trained physician adjudicators using hospital records, autopsy or coroner’s reports, or death certificates. Periodic linkage to the National Death Index was performed to verify deaths among participants who were lost to follow-up or whose death certificates or medical records were unavailable. Cause of death was centrally adjudicated.^23,24^ We examined only dementia-free deaths to rule out death due to dementia as a potential explanation of our findings.

We next examined the following composite outcome as our primary outcome of interest, which consisted of 3 categories: (1) survived to age 90 without cognitive impairment; (2) survived to age 90 with cognitive impairment (including mild cognitive impairment [MCI] and probable dementia); or (3) dementia-free death before age 90 (reference category). MCI and probable dementia were adjudicated annually through November 3, 2021.

The detailed WHIMS protocol for detecting MCI and probable dementia is described elsewhere.^19^ Briefly, in the main WHIMS trial, participants completed the Modified Mini-Mental State Examination, with those scoring below education-specific cutpoints (<80 for those with ≤8 years of education and <88 for those with ≥9 years of education) undergoing additional neuropsychological and standardized tests in person. A physician performed a neuropsychiatric evaluation with optional computerized tomography (without contrast) and blood assay and a friend or family member designated by the participant was interviewed regarding functional changes, classifying participants as having no dementia, MCI (based on Petersen’s criteria), or probable dementia (based on *Diagnostic and Statistical Manual of Mental Disorders, Fourth Edition* criteria).^25,26^ All data were reviewed by the WHIMS Clinical Coordinating Center, and final classification of cognitive status was centrally adjudicated by an adjudication panel consisting of a neurologist, geriatric psychiatrist, and geropsychologist. WHIMS-ECHO used a common and validated telephone-based cognitive assessment protocol with informant reviews and maintained similar procedures for ascertainment and central adjudication of final diagnosis.^22^

To classify cognitive impairment status at age 90, we prioritized WHIMS cognitive assessments within 3 years of the 90^th^ birth year. For participants who survived to age 90 but did not have a WHIMS cognitive assessment within three years of their 90^th^ birthday (N = 312), we determined cognitive impairment status at age 90 based on questionnaires administered to all WHI participants at baseline, 1-year, and 3-year follow-up visits, and annually after 2005. This questionnaire ascertained self-reported moderate or severe memory problems or physician-diagnosed dementia or AD; women were classified as having cognitive impairment if either of these conditions was reported. This approach was used in previous WHI analysis to define cognitive impairment status.^27^

### Covariates

Baseline questionnaires assessed age, race, ethnicity, education, smoking status, treated diabetes, cardiovascular disease (CVD, including myocardial infarction and stroke), non-melanoma cancer, and total physical activity summarized into metabolic equivalent (MET) hours per week based on duration, frequency, and intensity of walking and other recreational activities.^28^ Hypertension was defined as either self-report of physician-diagnosed hypertension, use of hypertensive medications, or measured systolic blood pressure ≥130 mm Hg or diastolic blood pressure ≥80 mm Hg. Height and weight were measured with a stadiometer and balance beam scale, respectively, to calculate body mass index (BMI; kg/m^2^). APOE ε2 and ε4 carrier status, defined as presence of at least one ε2 or ε4 allele, respectively, versus a non-carrier, was determined in women with available genome-wide genotyping data. Randomized assignment to hormone therapy treatment arm in the original WHIMS trial (estrogen alone, estrogen-alone placebo, estrogen plus progestin, or estrogen plus progestin placebo) was also included as a covariate. Additional covariates included high density lipoprotein (HDL) cholesterol, total cholesterol, and estimated glomerular filtration rate (eGFR), calculated using the 2021 Chronic Kidney Disease Epidemiology Collaboration (CKD-EPI) equation based on serum creatinine.^29^

### Statistical Methods

All plasma ADRD biomarkers were log_2_-transformed and subsequently standardized using z-scores to account for non-normal distributions and to allow comparisons with existing literature.^14^ To minimize the influence of outliers, we applied winsorization at the 99^th^ percentile across all plasma biomarkers. Means and standard deviations (SDs), or counts and proportions, were reported for continuous and categorical covariates, respectively, across quartiles of p-tau217, which was the biomarker we were most interested in due its strong associations with cognitive outcomes in prior studies, including WHIMS.^7,30,31^ Differences in continuous and categorical variables across p-tau217 quartiles were examined using Kruskal-Wallis rank sum tests and Chi-square tests or Fisher’s exact tests, respectively.

Logistic regression models were used to estimate odds ratios (ORs) and their 95% confidence intervals (CIs) for associations of one SD increases in log_2_-transformed plasma biomarkers with the odds of achieving exceptional longevity, defined as survival to age 90 vs dementia-free death before age 90. Subsequently, multinomial logistic regression models were used to estimate associations of plasma biomarkers with the composite outcome, comparing (1) survival to age 90 with cognitive impairment (MCI or probable dementia) and (2) survival to age 90 without cognitive impairment with (3) dementia-free death before age 90.

We examined four progressively adjusted models. Model 1 adjusted for age alone. Model 2 adjusted for age, race, and ethnicity. Model 3 adjusted for age, race, ethnicity, and hormone therapy trial arm. Model 4 further adjusted for education, smoking, physical activity, BMI, diabetes, CVD, cancer, eGFR, total cholesterol, HDL cholesterol, and hypertension. These potential confounders were selected from prior literature and include factors that may influence levels of plasma AD biomarkers (e.g., eGFR, BMI, co-morbidities, cholesterol levels).^14,32^ The *APOE* ε2 allele is associated with longevity while the *APOE* ε4 allele is a strong genetic risk factor for AD.^33^ APOE genotype was not included in our primary analyses because it was available only among White women; however, it was examined in subgroup analyses. Weights were generated to account for the sampling design, defined as the product of inverse propensity score weights (IPW) and sampling weights (see Supplementary Methods). Covariates with missing data were imputed using multivariate imputation by chained equations with 20 imputations and 20 iterations using the *mice* package in R.

Several sensitivity analyses were performed. We limited the sample to those who survived to age 90 to determine whether biomarkers could distinguish those who developed cognitive impairment from those who remained cognitively healthy at age 90. We examined whether findings varied by race (Black vs White); participants from underrepresented groups other than Black or White were excluded because of insufficient sample size. We also examined whether findings varied by *APOE* ε2 or *APOE* ε4 carrier status or hormone therapy (HT) assignment; interactions between plasma biomarkers and these factors were assessed using likelihood ratio tests. To assess the robustness of our findings against extreme plasma ADRD biomarker values, we repeated the analyses after removing outliers exceeding 5-SD above the mean instead of winsorizing outliers. Finally, we repeated the primary analyses including dementia-related deaths before age 90 (N = 181). All sensitivity analyses were based on the fully adjusted model.

Analyses were conducted using R 4.4.3 (https://www.r-project.org/) in RStudio 2024.12.1 (https://cran.rstudio.com/). We applied a Bonferroni-corrected threshold of p ≤ 0.005 (0.05/10) for significance to account for five biomarker associations across two outcomes (i.e., exceptional longevity and the composite outcome).

## RESULTS

### Baseline Characteristics

The analytic sample included 2,576 WHIMS participants. Differences between WHIMS participants in our analytic sample and those not included in the sample are reported in **Table S1**. In the analytic sample, the mean (SD) age was 70.0 (3.8) years; 72.5% were White, 18.9% were Black, 4.8% were Asian, 0.3% were Native Hawaiian or other Pacific Islander, 2.8% were more than one race, and 7.4% were Hispanic or Latino (**Table 1**). Higher plasma p-tau217 was associated with lower BMI, a higher prevalence of never smokers, a higher prevalence of non-melanoma cancer, lower eGFR, and a greater proportion of APOE ε4 carriers. The proportion of White women was larger at higher levels of plasma p-tau217, while the proportion of Black women was smaller. There were no significant differences by ethnicity, physical activity, education, diabetes, CVD, hypertension, total cholesterol, HDL cholesterol, or APOE ε2 carriage by plasma p-tau217.

**Table 1.**
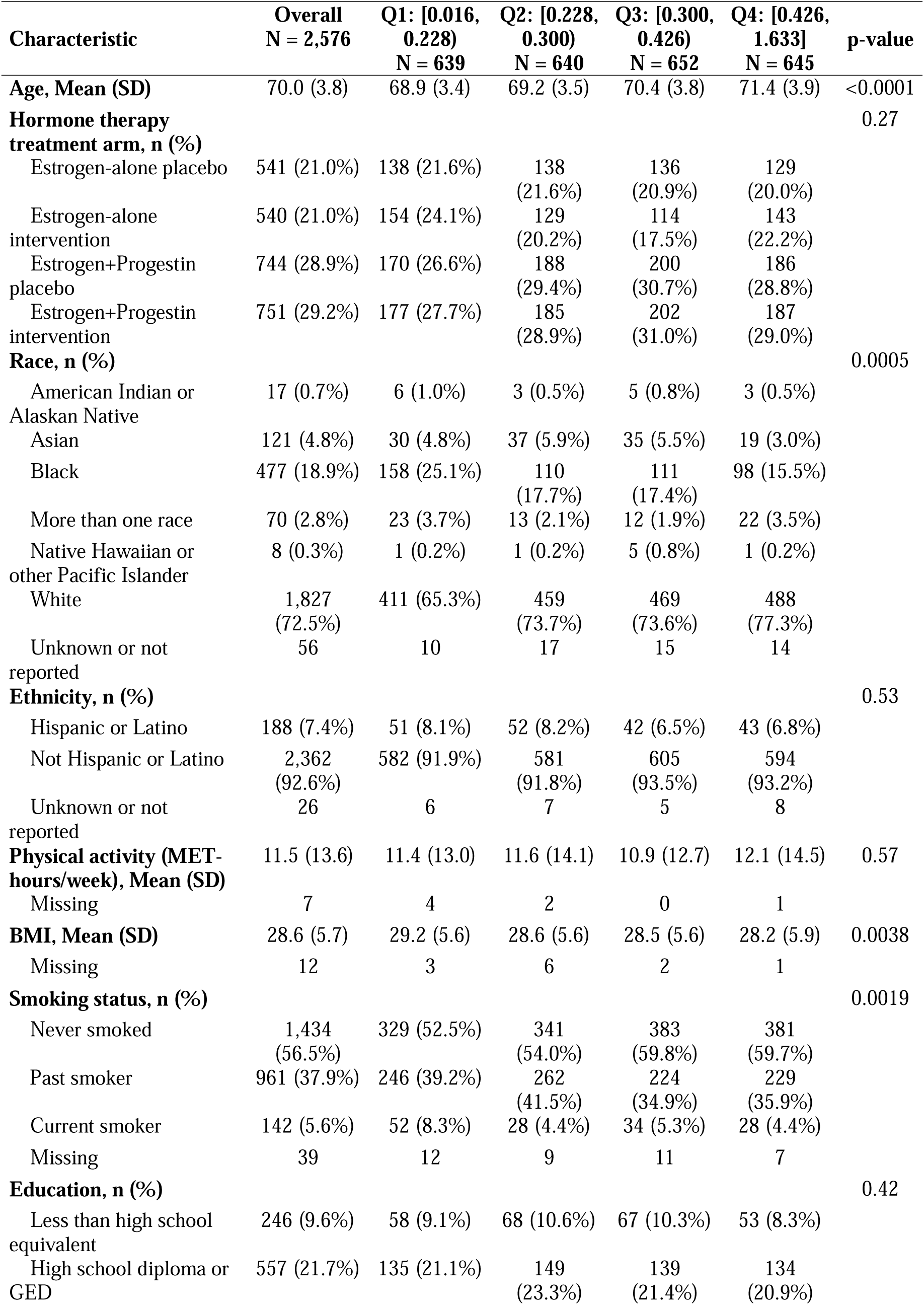
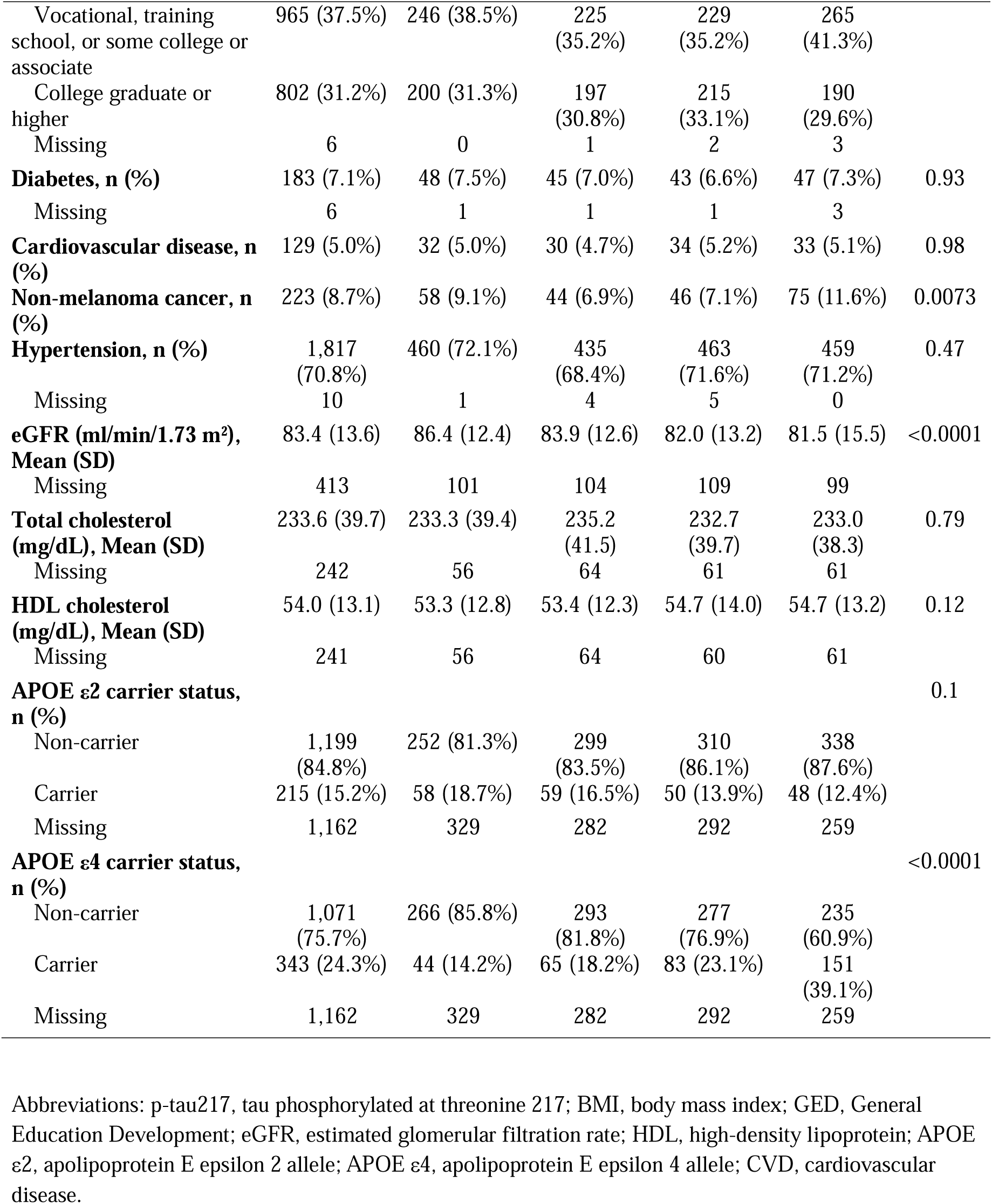
Baseline Characteristics by Quartiles of Plasma P-tau217.

The analytic sample included 1774 women who survived to age 90 during follow-up. Among the sample limited to those who had data on cognitive impairment status within 3 years of the 90^th^ birth year or who had dementia-free death before age 90, 32.6% survived to age 90 with cognitive impairment, and 33.9% survived to age 90 without cognitive impairment. Average levels of plasma ADRD biomarkers are shown in **Figure S2**. Women who survived to age 90 without cognitive impairment had lower baseline levels of plasma ADRD biomarkers than women who died before age 90 or survived to age 90 with cognitive impairment. Baseline biomarker levels were similar between women who died before age 90 or survived to age 90 with cognitive impairment.

### Survival to Age 90

In the fully adjusted models, elevated levels of p-tau217, NfL, and GFAP were associated with lower odds of survival to age 90 compared with dementia-free death before age 90. Only p-tau217 and NfL survived correction for multiple comparisons, with p-tau217 showing the largest magnitude of association (OR, 0.70; 95% CI, 0.61 – 0.81; p < 0.0001; **Figure 1, Table S2**).

**Figure 1.**
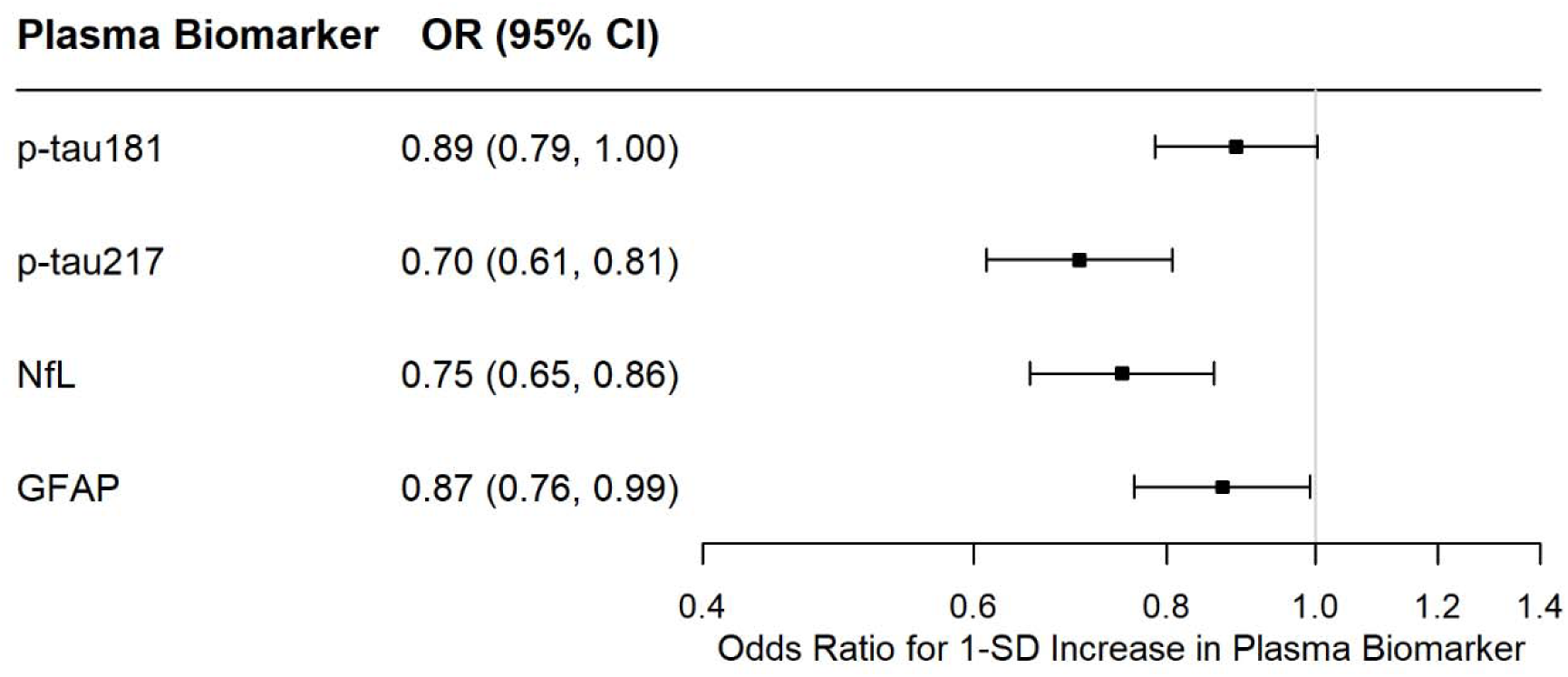
Associations of Plasma AD Biomarkers with Survival to Age 90 Versus Dementia-Free Death before Age 90. Abbreviations: AD, Alzheimer’s disease; OR, odds ratio; CI, confidence intervals; p-tau181, tau phosphorylated at threonine 181; p-tau217, tau phosphorylated at threonine 217; NfL = neurofilament light; GFAP = glial fibrillary acidic protein. a. Participants with missing race or ethnicity variables were excluded, leaving a final sample of N = 2,495. b. Results represent 1-SD increase in the log2 of plasma biomarker: p-tau181 (SD 0.54), p-tau217 (SD 0.78), NfL (SD 0.58), and GFAP (SD 0.63). c. The reference group was dementia-free death before age 90 (N = 761). d. Models adjusted for the following baseline covariates: age, hormone therapy arm, race, ethnicity, education, smoking, physical activity, BMI, diabetes, cardiovascular disease, non-melanoma cancer, eGFR, total cholesterol, HDL cholesterol, and hypertension. e. All models incorporated the product of inverse propensity score weights and sampling weights. f. Participants with missing race or ethnicity variables were excluded in models.

### Survival to Age 90 with or without Cognitive Impairment

In the fully adjusted models, all plasma biomarkers were associated with lower odds of surviving to age 90 without cognitive impairment compared with dementia-free death before age 90, with the largest magnitude of association observed for plasma p-tau217 (OR, 0.58; 95% CI, 0.50 – 0.68; p < 0.0001; **Figure 2, Table S3**). These associations survived correction for multiple comparisons. No plasma biomarker was associated with surviving to age 90 with cognitive impairment when compared with dementia-free death before age 90.

**Figure 2.**
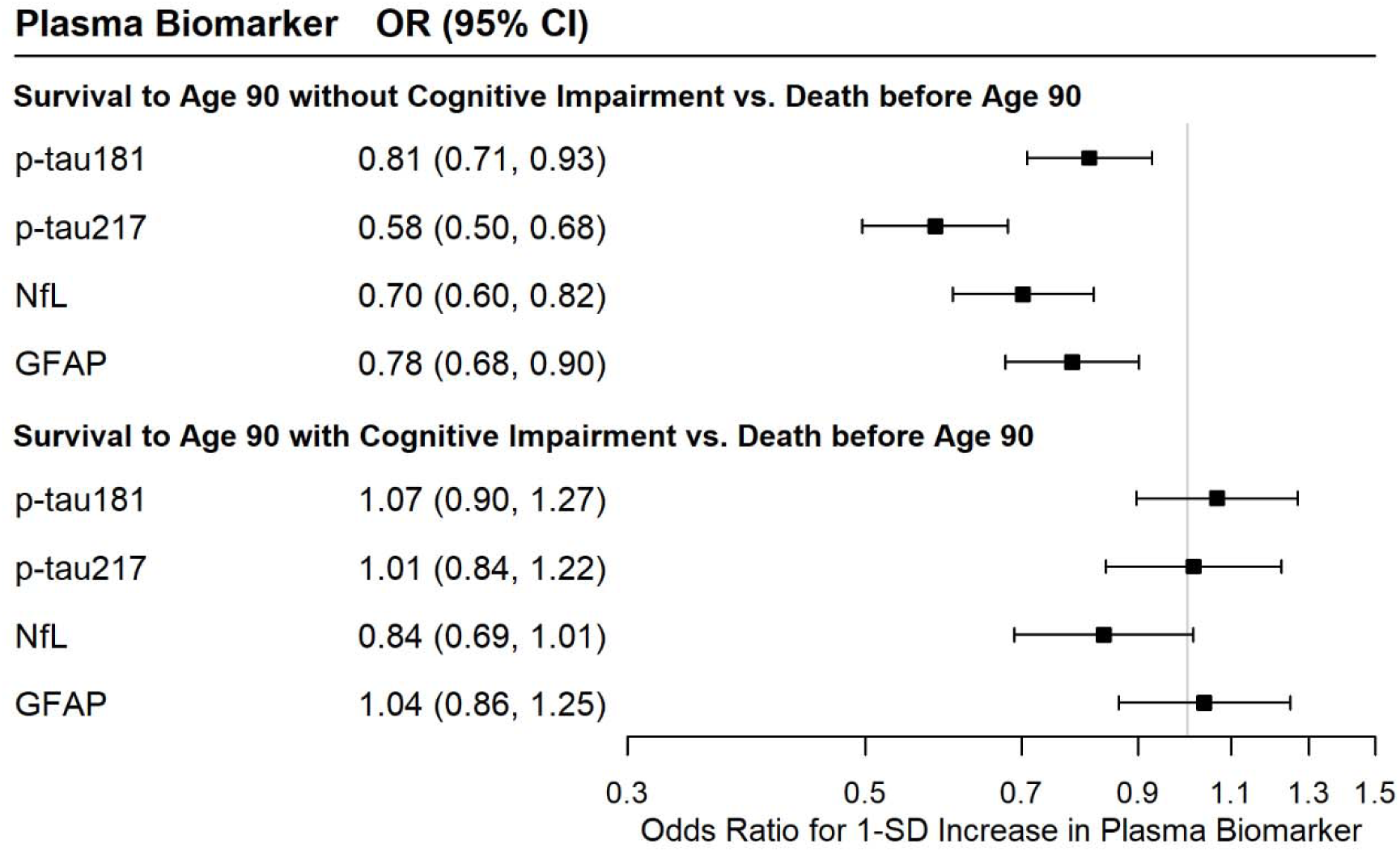
Associations of Plasma AD Biomarkers with Survival to Age 90 With and Without Cognitive Impairment Versus Dementia-Free Death before Age 90. Abbreviations: AD, Alzheimer’s disease; OR, odds ratio; CI, confidence intervals; p-tau181, tau phosphorylated at threonine 181; p-tau217, tau phosphorylated at threonine 217; NfL, neurofilament light; GFAP, glial fibrillary acidic protein. a. Participants with missing cognitively healthy longevity outcome, race, or ethnicity variables were excluded, leaving a final sample of N = 2,334. b. Results represent 1-SD increase in the log2 of plasma biomarker: p-tau181 (SD 0.54), p-tau217 (SD 0.78), NfL (SD 0.58), and GFAP (SD 0.63). c. The reference group was women who did not survive to age 90 years (N = 761). d. Models adjusted for the following baseline covariates: age, hormone therapy arm, race, ethnicity, education, smoking, physical activity, BMI, diabetes, cardiovascular disease, non-melanoma cancer, eGFR, total cholesterol, HDL cholesterol, and hypertension. e. All models incorporated the product of inverse propensity score weights and sampling weights. f. Participants with missing race or ethnicity variables were excluded in models.

### Sensitivity Analyses

When restricting analyses to those who survived to age 90, the odds of surviving to age 90 without cognitive impairment versus with cognitive impairment were lower for higher levels of plasma p-tau181 (OR, 0.76; 95% CI, 0.65 – 0.90), p-tau217 (OR, 0.55; 95% CI, 0.45 – 0.67), NfL (OR, 0.82; 95% CI, 0.68 – 0.99), and GFAP (OR, 0.74; 95% CI, 0.62 – 0.89) (**Figure 3, Table S4**).

**Figure 3.**
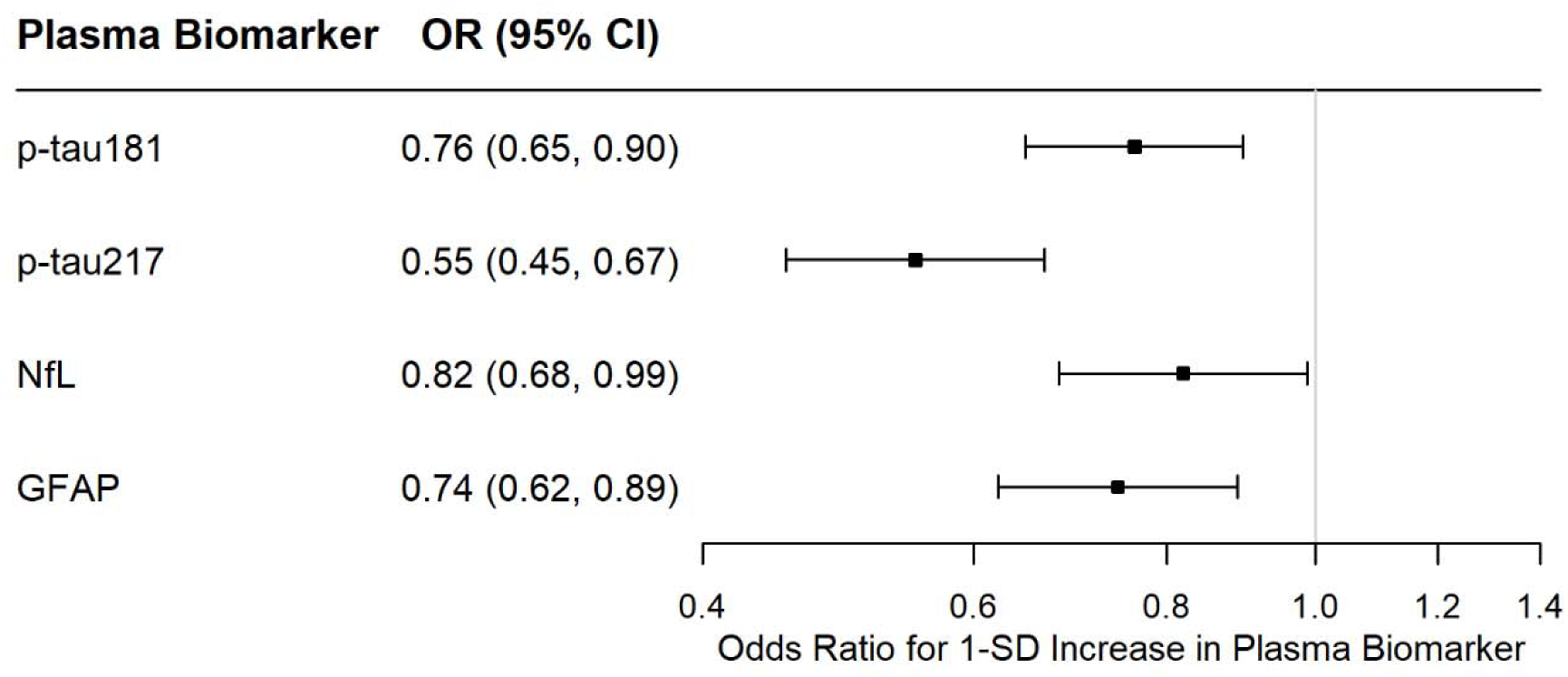
Associations of Plasma AD Biomarkers with Survival to Age 90 without Cognitive Impairment Versus Survival to Age 90 with Cognitive Impairment. Abbreviations: AD, Alzheimer’s disease; OR, odds ratio; CI, confidence intervals; p-tau181, tau phosphorylated at threonine 181; p-tau217, tau phosphorylated at threonine 217; NfL = neurofilament light; GFAP = glial fibrillary acidic protein. a. Participants with missing race or ethnicity variables were excluded, leaving a final analytic sample of N = 1,573. b. Results represent 1-SD increase in the log2 of plasma biomarker: p-tau181 (SD 0.54), p-tau217 (SD 0.78), NfL (SD 0.58), and GFAP (SD 0.63). a. The reference group was women who survived to age 90 with cognitive impairment (N = 772). b. Models adjusted for the following baseline covariates: age, hormone therapy arm, race, ethnicity, education, smoking, physical activity, BMI, diabetes, cardiovascular disease, non-melanoma cancer, eGFR, total cholesterol, HDL cholesterol, and hypertension. c. All models incorporated the product of inverse propensity score weights and sampling weights. d. Participants with missing race or ethnicity variables were excluded in models.

Associations did not vary by race, APOE ε2 or ε4 carriage, or by hormone therapy assignment (**Table S5-S9**). In sensitivity analyses excluding outliers defined as 5-SD above the mean and analyses including dementia deaths before age 90, findings were similar to the main analysis (**Table S10 and S11**).

## DISCUSSION

Higher levels of all plasma ADRD biomarkers at baseline were associated with lower odds of surviving to age 90 without cognitive impairment than dementia-free death before age 90 among older women. Plasma p-tau217 showed the largest association with cognitively healthy longevity, with each one standard deviation increase associated with 42% reduced odds of this outcome. Baseline biomarker levels did not distinguish between likelihood of attaining age 90 with cognitive impairment versus dementia-free death before age 90. These findings suggest that plasma ADRD biomarkers may serve as indicators of healthspan and identify those less likely to be cognitively healthy at the milestone age of 90 years.

A novel finding to emerge from our analysis is that plasma ADRD biomarkers may also represent biomarkers of aging. The most commonly examined biomarkers of aging have been omics-based biomarkers (e.g, epigenetic clocks) or individual protein molecules (e.g., interleukin-6).^34^ Such biomarkers of aging have been associated with age-related diseases, mortality, clinical phenotypes (e.g., frailty, cognitive function, walking speed), and healthspan, defined as the period of life in good health without chronic disease and disability.^34^ Plasma ADRD biomarkers have been previously associated with other age-related phenotypes, such as physical performance, gait speed, and mobility.^35–38^ Collectively, these results indicate that plasma ADRD biomarkers may predict diverse age-related phenotypes, including longevity and healthspan.

Women with elevated levels of plasma p-tau217 and NfL had reduced odds of surviving to age 90 than dying earlier from non-dementia causes. Higher levels of these biomarkers, and of p-tau181 and GFAP, were further associated with reduced odds of survival to age 90 without cognitive impairment. It is possible that biological processes captured by plasma ADRD biomarkers may extend beyond disease-specific mechanisms and may also reflect broader aging-related changes. Indeed, we recently observed that faster pace of epigenetic aging was associated with faster longitudinal increases in plasma p-tau217, p-tau181, GFAP, and NfL in WHIMS.^39^ These results suggest a potential link between biological aging and changes in Alzheimer’s pathology. Further studies are needed to determine the mechanisms underlying the associations between plasma ADRD biomarkers, longevity, and healthspan, including the potential role of biological aging.

This study has several strengths. WHIMS is a large, diverse cohort with information on a robust set of potential confounders, including demographic characteristics, lifestyle behaviors, and comorbidities. The long follow-up time to late life in the WHI allowed us to determine cognitive impairment status at the age of 90. MCI and ADRD were adjudicated through a comprehensive process involving neurocognitive assessments and final adjudication by a panel with expertise in dementia diagnosis. We had information on a comprehensive set of plasma ADRD biomarkers.

We also acknowledge several limitations. The sample was limited to older women. Findings need to be replicated in men and other cohorts with diverse racial and ethnic groups. Cognitively intact women were recruited at ages 65 and older, which likely explains the high survival rate to age 90 of 69% in our sample, conditional on their already advanced age at enrollment. The smaller sample size of Black women versus White women was also a limitation. APOE data were available only for White women.

In conclusion, we found that elevated levels of plasma biomarkers of ADRD pathology were associated with reduced odds of exceptional longevity and cognitively healthy longevity compared with dementia-free death. These findings suggest that plasma biomarkers may not only reflect ADRD pathology but also systemic aging processes that underlie lifespan and healthspan, underscoring their potential value as novel biomarkers of aging and as surrogate endpoints in anti-aging interventions.

## Data Availability

Data sharing statement
De-identified data from the study and supporting documents can be made available after publication to researchers with investigator support, after approval of a proposal by the WHI Publications and Presentations Committee and with a signed data access agreement (see https://www.whi.org/doc/PP-policy.pdf). Please contact Aladdin Shadyab and Linda McEvoy.

## Data sharing statement

De-identified data from the study and supporting documents can be made available after publication to researchers with investigator support, after approval of a proposal by the WHI Publications and Presentations Committee and with a signed data access agreement (see https://www.whi.org/doc/PP-policy.pdf). Please contact Aladdin Shadyab and Linda McEvoy.

## REFERENCES

1. Arias, E. United States life tables, 2000. Natl Vital Stat Rep 51, 1–38 (2002).

2. U.S. Census Bureau. The Older Population: 2020. https://www2.census.gov/library/publications/decennial/2020/census-briefs/c2020br-07.pdf (2023).

3. He, W. & Muenchrath, M. N. 90+ in the United States: 2006-2008. https://www2.census.gov/library/publications/2011/acs/acs-17.pdf (2011).

4. 2025 Alzheimer’s disease facts and figures. Alzheimer’s &amp; Dementia 21, (2025).

5. Hansson, O. et al. The Alzheimer’s Association appropriate use recommendations for blood biomarkers in Alzheimer’s disease. Alzheimers Dement 18, 2669–2686 (2022).

6. Janelidze, S. et al. Plasma P-tau181 in Alzheimer’s disease: relationship to other biomarkers, differential diagnosis, neuropathology and longitudinal progression to Alzheimer’s dementia. Nat Med 26, 379–386 (2020).

7. Lai, R., Li, B. & Bishnoi, R. P-tau217 as a Reliable Blood-Based Marker of Alzheimer’s Disease. Biomedicines 12, 1836 (2024).

8. Chatterjee, P. et al. Plasma glial fibrillary acidic protein is elevated in cognitively normal older adults at risk of Alzheimer’s disease. Transl Psychiatry 11, 27 (2021).

9. Dominantly Inherited Alzheimer Network et al. Serum neurofilament dynamics predicts neurodegeneration and clinical progression in presymptomatic Alzheimer’s disease. Nat Med 25, 277–283 (2019).

10. De Wolf, F. et al. Plasma tau, neurofilament light chain and amyloid-β levels and risk of dementia; a population-based cohort study. Brain 143, 1220–1232 (2020).

11. Verberk, I. M. W. et al. Combination of plasma amyloid beta(1-42/1-40) and glial fibrillary acidic protein strongly associates with cerebral amyloid pathology. Alz Res Therapy 12, 118 (2020).

12. Saji, N. et al. Relationship Between Plasma Neurofilament Light Chain, Gut Microbiota, and Dementia: A Cross-Sectional Study. J Alzheimers Dis 86, 1323–1335 (2022).

13. Cicognola, C. et al. Plasma glial fibrillary acidic protein detects Alzheimer pathology and predicts future conversion to Alzheimer dementia in patients with mild cognitive impairment. Alzheimers Res Ther 13, 68 (2021).

14. Lu, Y. et al. Changes in Alzheimer Disease Blood Biomarkers and Associations With Incident All-Cause Dementia. JAMA 332, 1258–1269 (2024).

15. Ferreira, P. C. L. et al. Plasma biomarkers identify older adults at risk of Alzheimer’s disease and related dementias in a real-world population-based cohort. Alzheimers Dement 19, 4507–4519 (2023).

16. Planche, V. et al. Validity and Performance of Blood Biomarkers for Alzheimer Disease to Predict Dementia Risk in a Large Clinic-Based Cohort. Neurology 100, e473–e484 (2023).

17. Martino-Adami, P. V. et al. Prognostic value of Alzheimer’s disease plasma biomarkers in the oldest-old: a prospective primary care-based study. Lancet Reg Health Eur 45, 101030 (2024).

18. Bullain, S. S. & Corrada, M. M. Dementia in the oldest old. Continuum (Minneap Minn) 19, 457–469 (2013).

19. Shumaker, S. A. et al. The Women’s Health Initiative Memory Study (WHIMS): a trial of the effect of estrogen therapy in preventing and slowing the progression of dementia. Control Clin Trials 19, 604–621 (1998).

20. Shumaker, S. A. et al. Estrogen Plus Progestin and the Incidence of Dementia and Mild Cognitive Impairment in Postmenopausal Women: The Women’s Health Initiative Memory Study: A Randomized Controlled Trial. JAMA 289, 2651 (2003).

21. Shumaker, S. A. Conjugated Equine Estrogens and Incidence of Probable Dementia and Mild Cognitive Impairment in Postmenopausal WomenWomen’s Health Initiative Memory Study. JAMA 291, 2947 (2004).

22. Espeland, M. A. et al. Long-term Effects on Cognitive Trajectories of Postmenopausal Hormone Therapy in Two Age Groups. GERONA glw156 (2016) doi:10.1093/gerona/glw156.

23. Curb, J. D. et al. Outcomes ascertainment and adjudication methods in the Women’s Health Initiative. Ann Epidemiol 13, S122–128 (2003).

24. Kabat, G. C. et al. White Blood Cell Count and Total and Cause-Specific Mortality in the Women’s Health Initiative. Am J Epidemiol 186, 63–72 (2017).

25. Petersen, R. C. et al. Current concepts in mild cognitive impairment. Arch Neurol 58, 1985–1992 (2001).

26. Bell, C. C. DSM-IV: Diagnostic and Statistical Manual of Mental Disorders. JAMA 272, 828 (1994).

27. Jain, P. et al. Analysis of Epigenetic Age Acceleration and Healthy Longevity Among Older US Women. JAMA Netw Open 5, e2223285 (2022).

28. Ainsworth, B. E. et al. Compendium of physical activities: classification of energy costs of human physical activities. Med Sci Sports Exerc 25, 71–80 (1993).

29. Inker, L. A. et al. New Creatinine- and Cystatin C-Based Equations to Estimate GFR without Race. N Engl J Med 385, 1737–1749 (2021).

30. Mendes, A. J. et al. Head-to-head study of diagnostic accuracy of plasma and cerebrospinal fluid p-tau217 versus p-tau181 and p-tau231 in a memory clinic cohort. J Neurol 271, 2053–2066 (2024).

31. Shadyab, A. H. et al. Plasma p-tau217 and incident mild cognitive impairment and dementia in older women: 25-year prospective study in The Women’s Health Initiative Memory Study. Preprint at 10.1101/2025.10.30.25339146 (2025).

32. Nguyen, S. et al. Associations of Epigenetic Age Estimators With Cognitive Function Trajectories in the Women’s Health Initiative Memory Study. Neurology 103, e209534 (2024).

33. Deelen, J. et al. A meta-analysis of genome-wide association studies identifies multiple longevity genes. Nat Commun 10, 3669 (2019).

34. Moqri, M. et al. Biomarkers of aging for the identification and evaluation of longevity interventions. Cell 186, 3758–3775 (2023).

35. Thompson, A. C. et al. Relationship of Alzheimer’s Disease and Related Dementias Plasma Biomarkers With Mobility in Cognitively Unimpaired Older Adults. J Gerontol A Biol Sci Med Sci 80, glaf110 (2025).

36. O’Bryant, S. E. et al. Plasma Biomarkers of Alzheimer’s Disease Are Associated with Physical Functioning Outcomes Among Cognitively Normal Adults in the Multiethnic HABS-HD Cohort. J Gerontol A Biol Sci Med Sci 78, 9–15 (2023).

37. He, L. et al. Cross-Sectional and Longitudinal Associations Between Plasma Neurodegenerative Biomarkers and Physical Performance Among Community-Dwelling Older Adults. J Gerontol A Biol Sci Med Sci 76, 1874–1881 (2021).

38. Ornago, A. M. et al. Blood biomarkers of Alzheimer’s disease and 12-year muscle strength trajectories in community-dwelling older adults: a cohort study. Lancet Healthy Longev 6, 100715 (2025).

39. Zhang, B. et al. Epigenetic clocks and longitudinal plasma biomarkers of Alzheimer’s disease. Alzheimers Dement 21, e70983 (2025).

