## Supplemental Material for "Associations of plasma biomarkers of Alzheimer’s pathology with longevity and healthspan"

**Table of Contents**

**Supplementary Methods**

**Figure S1.** Study Flow Chart

**Table S1**. Baseline Sociodemographic, Behavioral, and Health Characteristics by Analytic Sample Inclusion Status

**Figure S2**. Plasma ADRD Biomarkers Levels by Cognitively Healthy Longevity Outcomes

**Table S2**. Associations of Plasma ADRD Biomarkers with Survival to Age 90 vs Dementia-free Death before Age 90

**Table S3.** Associations of Plasma ADRD Biomarkers with Survival to Age 90 With and Without Cognitive Impairment Versus Dementia-free Death Before Age 90.

**Table S4**. Associations of Plasma ADRD Biomarkers with Survival to Age 90 Without versus With Cognitive Impairment in Those Who Survived to Age 90.

**Table S5**. Associations of Plasma ADRD Biomarkers with Odds of Surviving to Age 90 With or Without Cognitive Impairment, Versus Dementia-free Death Before Age 90 in White vs Black Women.

**Table S6.** Associations of Plasma ADRD Biomarkers with Odds of Surviving to Age 90 With or Without Cognitive Impairment, Versus Dementia-free Death Before Age 90 Stratified by APOE ε2 Carrier Status.

**Table S7.** Associations of Plasma ADRD Biomarkers with Odds of Surviving to Age 90 With or Without Cognitive Impairment, Versus Dementia-free Death Before Age 90 Stratified by APOE ε4 Carrier Status

**Table S8.** Associations of Plasma ADRD Biomarkers with Odds of Surviving to Age 90 With or Without Cognitive Impairment, Versus Dementia-free Death Before Age 90 Stratified by Randomization to Estrogen Alone vs Placebo

**Table S9.** Associations of Plasma ADRD Biomarkers with Odds of Surviving to Age 90 With or Without Cognitive Impairment, Versus Dementia-free Death Before Age 90 Stratified by Randomization to Estrogen plus Progestin versus Placebo

**Table S10.** Associations of Plasma ADRD Biomarkers with Odds of Surviving to Age 90 With or Without Cognitive Impairment, Versus Dementia-free Death Before Age 90 Stratified After Removing Outliers (5-SD above the Mean)

**Table S11.** Associations of Plasma ADRD Biomarkers with Odds of Surviving to Age 90 With or Without Cognitive Impairment Versus Death Before Age 90, Including Dementia Deaths.

**WHI Protocol for Blood Collection and Processing**

https://www.whi.org/doc/Vol-2-11-Blood-and-Urine-Collection-Processing-and-Shipment.pdf

**Inter-assay laboratory coefficients**

The following inter-assay laboratory coefficients of variation (CVs) were derived from an ARDL pooled sample and the two kit controls, which were run on every plate along with the samples: 1) NfL: 6.4%, 9.0%, and 9.4% at mean concentrations of 23.5, 499.2, and 8.2 pg/mL, respectively; 2) GFAP: 10.6%, 9.8%, and 15.7% at mean concentrations of 188.4, 3744.0, and 72.3 pg/mL, respectively; 3) p-tau181: 7.2%, 6.4%, and 10.7% at mean concentrations of 42.4, 1130.9, and 11.9 pg/mL, respectively; and 4) p-tau217: 11.4%, 11.2%, and 12.9% at mean concentrations of 0.75, 0.39, and 0.15 pg/mL, respectively.

**Lower limits of detection (LOD) for each biomarker.**

The lower limits of detection (LOD) for each biomarker were as follows: 1.76 pg/ml for GFAP, 0.36 pg/ml for NfL, 0.620 pg/ml for ptau181, and 0.012 pg/ml for ptau217. No woman in our analytic sample had values less than the LOD for any biomarker.

**Sample Selection**

Among the 7,479 WHIMS participants, we first selected all 1,334 women with incident MCI or probable dementia through the end of follow-up on November 3, 2021. We next selected 1,502 controls who did not have MCI or probable dementia during follow-up, including 565 who were enrolled in WHIMS-ECHO. Controls in our sample included all women who had brain imaging data (n=519) and all women from underrepresented populations (including American Indian/Alaskan Native, Asian, Native Hawaiian/Other Pacific Islander, Black, more than one race, and Hispanic/Latina; n=707). We excluded participants with missing plasma biomarker data (N = 52), those born after Feb 17, 1934 (N = 24), and those who died before age 90 due to dementia (N = 184), leading to a final analytic sample of 2,576 women. The logistic regression models were focused on women without missing race or ethnicity (N=2,495), as these variables were not imputed in the analysis.

**Calculation of Weights**

To account for differences between the full WHIMS cohort and the biomarker analytic sample, we generated inverse probability weights (IPW) for selection into the biomarker sample. Propensity scores for IPW were estimated using logistic regression in the full WHIMS cohort (N=7,479) based on covariates that may influence selection into the sample. The model estimated the probability of inclusion in the biomarker sample as a function of age, region, race, ethnicity, baseline smoking status, hormone therapy trial arm, baseline cardiovascular disease, diabetes, cancer, depressive symptoms, hysterectomy, prior hormone use, body mass index, hypertension, MCI/dementia diagnosis, and participation in Women's Health Initiative (WHI) ancillary studies (i.e., brain imaging and the Long Life Study).

To account for oversampling of MCI/dementia cases in our analytic sample, we generated sampling weights so that the weighted analytic sample reflected the population incidence of MCI/probable dementia in WHIMS.[1,2] Final weights for analysis were calculated as the product of IPW and the sampling weights. All analyses incorporated these final weights to generate estimates that were generalizable to the full WHIMS cohort.

### Reference

1 van der Laan MJ. Estimation based on case-control designs with known prevalence probability. *Int J Biostat* 2008; **4**: Article 17.

2 Rose S, van der Laan MJ. A note on risk prediction for case-control studies. 2008.

Figure S1. Study Flow Chart

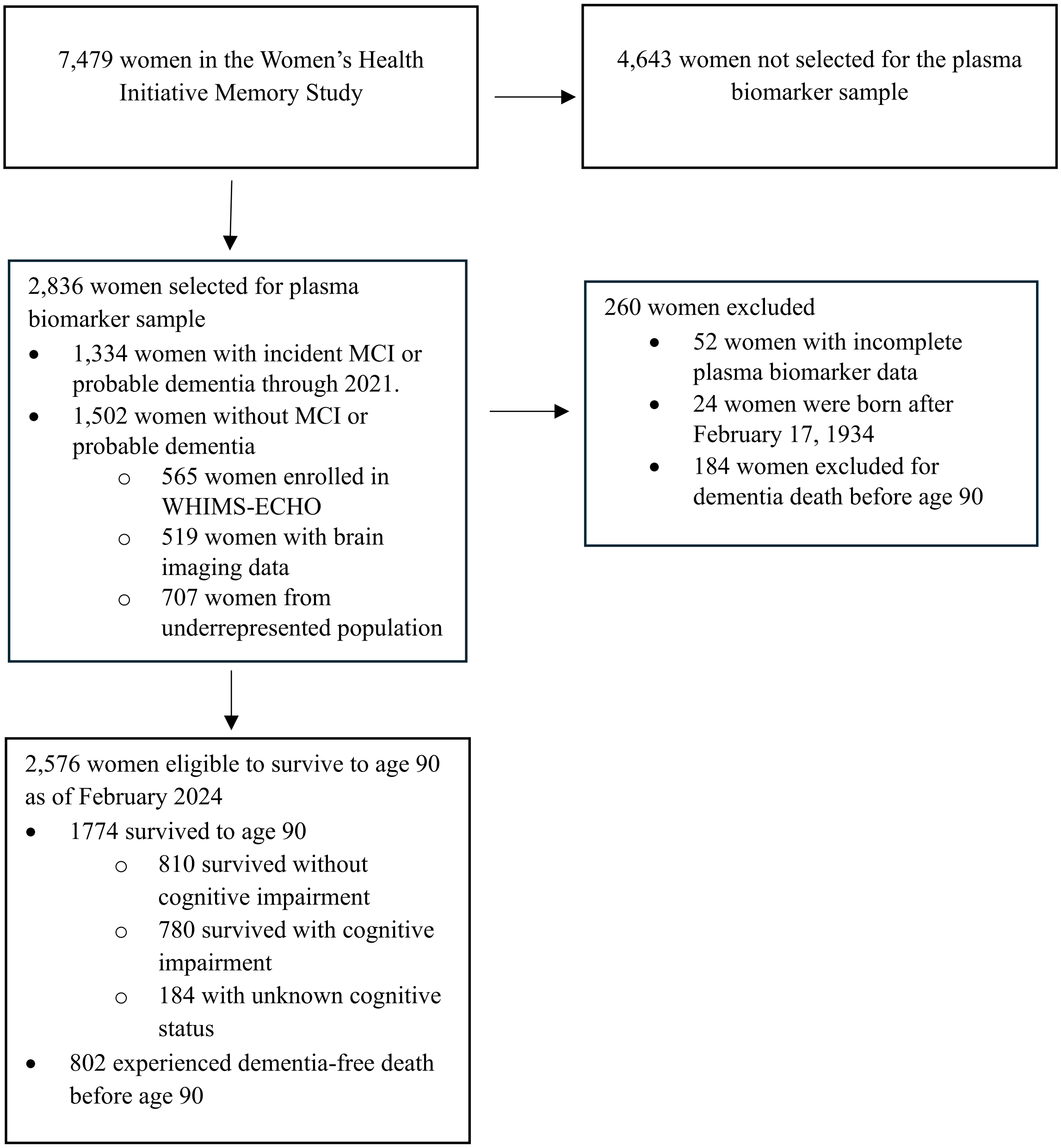

Table S1. Baseline Sociodemographic, Behavioral, and Health Characteristics by Analytic Sample Inclusion Status

| | Full WHIMS (N = 7,479) | Analytic Sample (N = 2,576) | Not in Analytic Sample (N = 4,903) | p-value |
|---|---|---|---|---|
| **Age, Mean (SD)** | 70.1 (3.8) | 70.0 (3.8) | 70.2 (3.9) | 0.019 |
| **Hormone Therapy Study Arm, n (%)** | | | | 0.011 |
| Estrogen-alone placebo | 1,484 (19.8%) | 541 (21.0%) | 943 (19.2%) | |
| Estrogen-alone intervention | 1,469 (19.6%) | 540 (21.0%) | 929 (18.9%) | |
| Estrogen + Progestin placebo | 2,302 (30.8%) | 744 (28.9%) | 1,558 (31.8%) | |
| Estrogen + Progestin intervention | 2,224 (29.7%) | 751 (29.2%) | 1,473 (30.0%) | |
| **Race, n (%)** | | | | 0.0005 |
| American Indian or Alaskan Native | 17 (0.2%) | 17 (0.7%) | 0 (0.0%) | |
| Asian | 127 (1.7%) | 121 (4.8%) | 6 (0.1%) | |
| Black | 523 (7.1%) | 477 (18.9%) | 46 (0.9%) | |
| More than one race | 80 (1.1%) | 70 (2.8%) | 10 (0.2%) | |
| Native Hawaiian or other Pacific Islander | 8 (0.1%) | 8 (0.3%) | 0 (0.0%) | |
| White | 6,649 (89.8%) | 1,827 (72.5%) | 4,822 (98.7%) | |
| Missing | 75 | 56 | 19 | |
| **Ethnicity, n (%)** | | | | <0.0001 |
| Hispanic or Latino | 214 (2.9%) | 188 (7.4%) | 26 (0.5%) | |
| Not Hispanic or Latino | 7,233 (97.1%) | 2,362 (92.6%) | 4,871 (99.5%) | |
| Missing | 32 | 26 | 6 | |
| **BMI, Mean (SD)** | 28.5 (5.7) | 28.6 (5.7) | 28.5 (5.7) | 0.16 |
| Missing | 43 | 12 | 31 | |
| **Smoking Status, n (%)** | | | | <0.0001 |
| Never Smoked | 3,909 (53.1%) | 1,434 (56.5%) | 2,475 (51.2%) | |
| Past Smoker | 2,930 (39.8%) | 961 (37.9%) | 1,969 (40.8%) | |
| Current Smoker | 529 (7.2%) | 142 (5.6%) | 387 (8.0%) | |
| Missing | 111 | 39 | 72 | |
| **Education, n (%)** | | | | <0.0001 |
| Less than high school equivalent | 576 (7.7%) | 246 (9.6%) | 330 (6.8%) | |
| High school diploma or GED | 1,647 (22.1%) | 557 (21.7%) | 1,090 (22.3%) | |
| Vocational, training school, or some college or associate | 3,002 (40.3%) | 965 (37.5%) | 2,037 (41.7%) | |
| College graduate or higher | 2,232 (29.9%) | 802 (31.2%) | 1,430 (29.3%) | |
| Missing | 22 | 6 | 16 | |
| **Diabetes, n (%)** | 488 (6.5%) | 183 (7.1%) | 305 (6.2%) | 0.14 |
| Missing | 14 | 6 | 8 | |
| **Cardiovascular disease, n (%)** | 366 (4.9%) | 129 (5.0%) | 237 (4.8%) | 0.74 |
| **Physical activity (hours/week), Mean (SD)** | 11.3 (13.3) | 11.5 (13.6) | 11.1 (13.1) | 0.32 |

| | | | | |
|---|---|---|---|---|
| Missing | 17 | 7 | 10 | |
| **Total cholesterol (mg/dL), Mean (SD)** | 234.4 (40.0) | 234.3 (39.8) | 234.4 (40.1) | 0.98 |
| Missing | 985 | 412 | 573 | |
| **HDL cholesterol (mg/dL), Mean (SD)** | 53.4 (12.6) | 53.7 (12.6) | 53.3 (12.5) | 0.17 |
| Missing | 985 | 412 | 573 | |
| **Hypertension, n (%)** | 5,269 (70.6%) | 1,817 (70.8%) | 3,452 (70.6%) | 0.82 |
| Missing | 21 | 10 | 11 | |
| **eGFR (ml/min/1.73 m²), Mean (SD)** | 84.0 (13.5) | 83.4 (13.6) | 84.3 (13.4) | 0.0046 |
| Missing | 986 | 413 | 573 | |
| **APOE ε2 carrier status, n (%)** | | | | 0.86 |
| Non-carrier | 4,141 (84.6%) | 1,199 (84.8%) | 2,942 (84.6%) | |
| Carrier | 751 (15.4%) | 215 (15.2%) | 536 (15.4%) | |
| Missing | 2,587 | 1,162 | 1,425 | |
| **APOE ε4 carrier status, n (%)** | | | | 0.47 |
| Non-carrier | 3,671 (75.0%) | 1,071 (75.7%) | 2,600 (74.8%) | |
| Carrier | 1,221 (25.0%) | 343 (24.3%) | 878 (25.2%) | |
| Missing | 2,587 | 1,162 | 1,425 | |

Abbreviations: BMI, body mass index; HDL, high-density lipoprotein; eGFR, estimated glomerular filtration rate; APOE ε2, apolipoprotein E epsilon 2 allele; APOE ε4, apolipoprotein E epsilon 4

Figure S2. Plasma ADRD Biomarkers Levels by Cognitively Healthy Longevity Outcomes

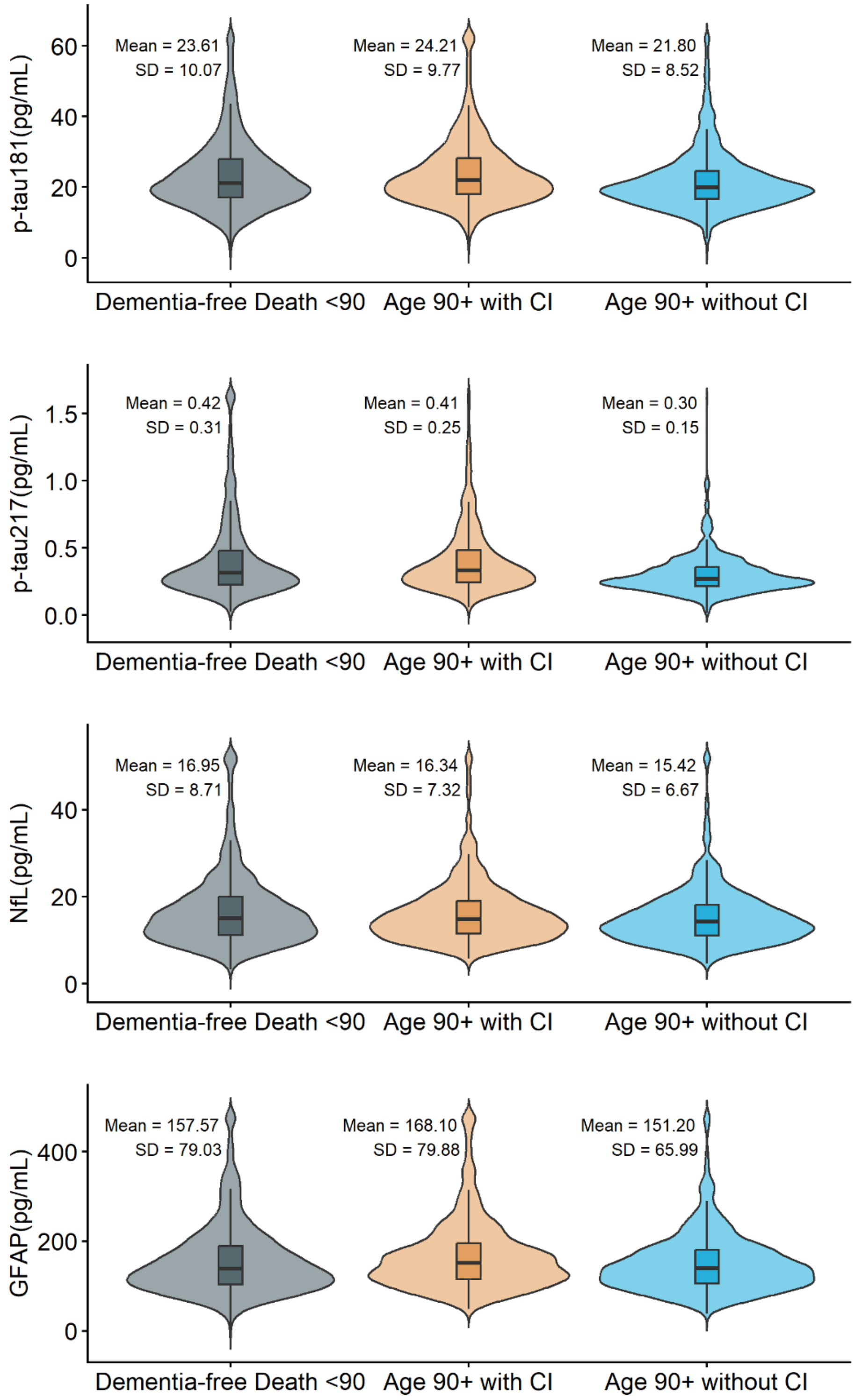


Abbreviations: ADRD, Alzheimer's disease and related dementias; CI cognitive impairment; SD, standard deviation; OR, odds ratio; CI, Cognitive Impairment; p-tau181, tau phosphorylated at threonine 181; p-tau217, tau phosphorylated at threonine 217, GFAP, glial fibrillary acidic protein.

Table S2. Associations of Plasma ADRD Biomarkers with Survival to Age 90 vs Dementia-free Death before Age 90

| Biomarker | SD | Model | OR (95% CI) | P-value |
|---|---|---|---|---|
| p-tau181 | 0.54 | 1 | 0.92 (0.82, 1.03) | 0.14 |
| | | 2 | 0.91 (0.81, 1.02) | 0.11 |
| | | 3 | 0.91 (0.81, 1.02) | 0.10 |
| | | 4 | 0.89 (0.79, 1.00) | 0.055 |
| p-tau217 | 0.78 | 1 | 0.77 (0.67, 0.87) | <0.0001 |
| | | 2 | 0.74 (0.65, 0.84) | <0.0001 |
| | | 3 | 0.74 (0.65, 0.84) | <0.0001 |
| | | 4 | 0.70 (0.61, 0.81) | <0.0001 |
| NfL | 0.58 | 1 | 0.81 (0.72, 0.91) | 0.0006 |
| | | 2 | 0.80 (0.70, 0.90) | 0.0003 |
| | | 3 | 0.79 (0.70, 0.90) | 0.0003 |
| | | 4 | 0.75 (0.65, 0.86) | <0.0001 |
| GFAP | 0.63 | 1 | 0.97 (0.86, 1.10) | 0.64 |
| | | 2 | 0.96 (0.85, 1.09) | 0.52 |
| | | 3 | 0.96 (0.85, 1.09) | 0.53 |
| | | 4 | 0.87 (0.76, 0.99) | 0.038 |

Abbreviations: ADRD, Alzheimer's disease and related dementias; SD; standard deviation; OR, odds ratio; CI, confidence intervals; p-tau181, tau phosphorylated at threonine 181; p-tau217, tau phosphorylated at threonine 217; NfL, neurofilament light; GFAP, glial fibrillary acidic protein.

a. Participants with missing race or ethnicity variables were excluded.
b. The reference group was dementia-free death before age 90 (N = 761).
c. Model 1 adjusted for age. Model 2 adjusted for age, race, and ethnicity. Model 3 adjusted for age, race, ethnicity, and hormone therapy trial arm. Model 4 adjusted for age, race, ethnicity, hormone therapy trial arm, education, smoking status, physical activity, BMI, diabetes, cardiovascular disease, non-melanoma cancer, estimated glomerular filtration rate, total cholesterol, high-density lipoprotein cholesterol, and hypertension.
d. Results represent 1-SD increase in the log2-transformed plasma biomarker. All models incorporated the product of inverse propensity score weights and sampling weights.

Table S3. Associations of Plasma ADRD Biomarkers with Survival to Age 90 With and Without Cognitive Impairment Versus Dementia-free Death Before Age 90.

| | | | Without Cognitive Impairment (N = 801) | | With Cognitive Impairment (N = 772) | |
|---|---|---|---|---|---|---|
| Biomarker | SD | Model | OR (95% CI) | P-value | OR (95% CI) | P-value |
| p-tau181 | 0.54 | 1 | 0.87 (0.77, 0.98) | 0.025 | 1.11 (0.94, 1.31) | 0.21 |
| | | 2 | 0.84 (0.74, 0.96) | 0.0077 | 1.09 (0.92, 1.28) | 0.33 |
| | | 3 | 0.84 (0.74, 0.95) | 0.0058 | 1.08 (0.92, 1.28) | 0.35 |
| | | 4 | 0.81 (0.71, 0.93) | 0.0021 | 1.07 (0.90, 1.27) | 0.47 |
| p-tau217 | 0.78 | 1 | 0.68 (0.59, 0.78) | <0.0001 | 1.10 (0.92, 1.32) | 0.31 |
| | | 2 | 0.62 (0.54, 0.72) | <0.0001 | 1.05 (0.87, 1.26) | 0.63 |
| | | 3 | 0.62 (0.54, 0.72) | <0.0001 | 1.05 (0.87, 1.26) | 0.63 |
| | | 4 | 0.58 (0.50, 0.68) | <0.0001 | 1.01 (0.84, 1.22) | 0.89 |
| NfL | 0.58 | 1 | 0.79 (0.70, 0.90) | 0.0004 | 0.88 (0.73, 1.05) | 0.14 |
| | | 2 | 0.77 (0.67, 0.88) | 0.0001 | 0.87 (0.72, 1.03) | 0.11 |
| | | 3 | 0.76 (0.67, 0.88) | 0.0001 | 0.86 (0.72, 1.03) | 0.11 |
| | | 4 | 0.70 (0.60, 0.82) | <0.0001 | 0.84 (0.69, 1.01) | 0.069 |
| GFAP | 0.63 | 1 | 0.91 (0.80, 1.04) | 0.15 | 1.15 (0.96, 1.38) | 0.12 |
| | | 2 | 0.88 (0.77, 1.00) | 0.055 | 1.13 (0.94, 1.35) | 0.20 |
| | | 3 | 0.87 (0.76, 1.00) | 0.051 | 1.13 (0.94, 1.35) | 0.20 |
| | | 4 | 0.78 (0.68, 0.90) | 0.0007 | 1.04 (0.86, 1.25) | 0.70 |

Abbreviations: ADRD, Alzheimer's disease and related dementias; SD; standard deviation; OR, odds ratio; CI, confidence intervals; p-tau181, tau phosphorylated at threonine 181; p-tau217, tau phosphorylated at threonine 217; NfL, neurofilament light; GFAP, glial fibrillary acidic protein.

a. Participants with missing cognitively healthy longevity outcome, race, or ethnicity variables were excluded.
b. The reference group was dementia-free death before age 90 (N = 761).
c. Model 1 adjusted for age. Model 2 adjusted for age, race, and ethnicity. Model 3 adjusted for age, race, ethnicity, and hormone therapy trial arm. Model 4 adjusted for age, race, ethnicity, hormone therapy trial arm, education, smoking status, physical activity, BMI, diabetes, cardiovascular disease, non-melanoma cancer, estimated glomerular filtration rate, total cholesterol, high-density lipoprotein cholesterol, and hypertension.
d. Results represent 1-SD increase in the log2-transformed plasma biomarker. All models incorporated the product of inverse propensity score weights and sampling weights.

Table S4. Associations of Plasma ADRD Biomarkers with Survival to Age 90 Without versus With Cognitive Impairment in Those Who Survived to Age 90.

| Biomarker | SD | Model | OR (95% CI) | P-Value |
|---|---|---|---|---|
| p-tau181 | 0.54 | 1 | 0.77 (0.66, 0.91) | 0.0015 |
| | | 2 | 0.78 (0.66, 0.91) | 0.0016 |
| | | 3 | 0.77 (0.66, 0.90) | 0.0014 |
| | | 4 | 0.76 (0.65, 0.90) | 0.0011 |
| p-tau217 | 0.78 | 1 | 0.58 (0.48, 0.70) | <0.0001 |
| | | 2 | 0.57 (0.47, 0.68) | <0.0001 |
| | | 3 | 0.57 (0.47, 0.68) | <0.0001 |
| | | 4 | 0.55 (0.45, 0.67) | <0.0001 |
| NfL | 0.58 | 1 | 0.90 (0.76, 1.06) | 0.22 |
| | | 2 | 0.87 (0.73, 1.03) | 0.11 |
| | | 3 | 0.87 (0.73, 1.03) | 0.11 |
| | | 4 | 0.82 (0.68, 0.99) | 0.036 |
| GFAP | 0.63 | 1 | 0.77 (0.65, 0.91) | 0.0029 |
| | | 2 | 0.76 (0.64, 0.90) | 0.0020 |
| | | 3 | 0.76 (0.64, 0.90) | 0.0020 |
| | | 4 | 0.74 (0.62, 0.89) | 0.0013 |

Abbreviations: ADRD, Alzheimer's disease and related dementias; SD; standard deviation; OR, odds ratio; CI, confidence intervals; p-tau181, tau phosphorylated at threonine 181; p-tau217, tau phosphorylated at threonine 217; NfL, neurofilament light; GFAP, glial fibrillary acidic protein.

a. Participants with missing race or ethnicity variables were excluded.
b. The reference group was women who survived to age 90 with cognitive impairment (N = 772).
c. Model 1 adjusted for age. Model 2 adjusted for age, race, and ethnicity. Model 3 adjusted for age, race, ethnicity, and hormone therapy trial arm. Model 4 adjusted for age, race, ethnicity, hormone therapy trial arm, education, smoking status, physical activity, BMI, diabetes, cardiovascular disease, non-melanoma cancer, estimated glomerular filtration rate, total cholesterol, high-density lipoprotein cholesterol, and hypertension.
d. Results represent 1-SD increase in the log2-transformed plasma biomarker. All models incorporated the product of inverse propensity score weights and sampling weights.

Table S5. Associations of Plasma ADRD Biomarkers with Odds of Surviving to Age 90 With or Without Cognitive Impairment, Versus Dementia-free Death Before Age 90 in White vs Black Women.

| | White | | | Black | | | |
|---|---|---|---|---|---|---|---|
| | | Without Cognitive Impairment (N = 645) | With Cognitive Impairment (N = 645) | | Without Cognitive Impairment (N = 100) | With Cognitive Impairment (N = 100) | |
| Biomarker | SD | OR (95% CI) | OR (95% CI) | SD | OR (95% CI) | OR (95% CI) | P-interaction |
| p-tau181 | 0.53 | 0.80 (0.68, 0.93) | 1.05 (0.86, 1.29) | 0.61 | 0.97 (0.70, 1.34) | 1.09 (0.72, 1.66) | 0.72 |
| p-tau217 | 0.78 | 0.55 (0.46, 0.66) | 0.95 (0.76, 1.18) | 0.80 | 0.72 (0.50, 1.02) | 1.18 (0.75, 1.84) | 0.85 |
| NfL | 0.56 | 0.70 (0.59, 0.83) | 0.86 (0.69, 1.07) | 0.66 | 0.52 (0.35, 0.77) | 0.64 (0.40, 1.04) | 0.085 |
| GFAP | 0.61 | 0.78 (0.66, 0.92) | 1.06 (0.85, 1.31) | 0.69 | 0.76 (0.55, 1.05) | 0.91 (0.59, 1.40) | 0.58 |

Abbreviations: ADRD, Alzheimer's disease and related dementias; SD, standard deviation; OR, odds ratio; CI, confidence intervals; p-tau181, tau phosphorylated at threonine 181; p-tau217, tau phosphorylated at threonine 217.

a. Participants with missing cognitively healthy longevity outcome, race, or ethnicity variables were excluded.
b. The reference group was dementia-free death before age 90 (White: N = 495; Black: N = 204).
c. Model adjusted for the following baseline covariates: age, hormone therapy arm, education, smoking, physical activity, BMI, diabetes, cardiovascular disease, non-melanoma cancer, estimated glomerular filtration rate, total cholesterol, high-density lipoprotein cholesterol, hypertension.
d. P-values for interactions were derived from likelihood ratio tests comparing nested models with and without the interaction term between plasma biomarker and race.
e. Results represent 1-SD increase in the log2 of plasma biomarkers. All models incorporated the product of inverse propensity score weights and sampling weights.

Table S6. Associations of Plasma ADRD Biomarkers with Odds of Surviving to Age 90 With or Without Cognitive Impairment, Versus Dementia-free Death Before Age 90 Stratified by APOE ε2 Carrier Status.

| | | APOE ε2 Non-carriers | | | APOE ε2 carriers | | |
|---|---|---|---|---|---|---|---|
| | | Without Cognitive Impairment (N = 432) | With Cognitive Impairment (N = 426) | | Without Cognitive Impairment (N = 88) | With Cognitive Impairment (N = 75) | |
| Biomarker | SD | OR (95% CI) | OR (95% CI) | SD | OR (95% CI) | OR (95 CI) | P-Interaction |
| p-tau181 | 0.53 | 0.80 (0.66, 0.97) | 1.09 (0.85, 1.40) | 0.48 | 0.72 (0.41, 1.25) | 0.68 (0.35, 1.33) | 0.18 |
| p-tau217 | 0.78 | 0.51 (0.41, 0.65) | 0.97 (0.73, 1.27) | 0.71 | 0.42 (0.22, 0.80) | 0.50 (0.22, 1.11) | 0.13 |
| NfL | 0.56 | 0.76 (0.61, 0.94) | 0.94 (0.71, 1.24) | 0.56 | 0.38 (0.18, 0.81) | 0.51 (0.20, 1.28) | 0.24 |
| GFAP | 0.62 | 0.75 (0.60, 0.93) | 1.12 (0.85, 1.46) | 0.60 | 0.42 (0.22, 0.79) | 0.91 (0.42, 1.96) | 0.32 |

Abbreviations: ADRD, Alzheimer's disease and related dementias; SD, standard deviation; OR, odds ratio; CI, confidence intervals; p-tau181, tau phosphorylated at threonine 181; p-tau217, tau phosphorylated at threonine 217, GFAP, glial fibrillary acidic protein.

a. Participants with missing cognitively healthy longevity outcome or APOE ε4 variables were excluded.
b. The reference group was dementia-free death before age 90 (Non-carrier: N = 323; carrier: N = 45).
c. Model adjusted for the following baseline covariates: age, hormone therapy arm, education, smoking, physical activity, BMI, diabetes, cardiovascular disease, non-melanoma cancer, estimated glomerular filtration rate, total cholesterol, high-density lipoprotein cholesterol, and, hypertension.
d. P-values for interactions were derived from likelihood ratio tests comparing nested models with and without the interaction term between plasma biomarker and APOE ε4.
e. Results represent 1-SD increase in the log2 of plasma biomarkers. All models incorporated the product of inverse propensity score weights and sampling weights.

Table S7. Associations of Plasma ADRD Biomarkers with Odds of Surviving to Age 90 With or Without Cognitive Impairment, Versus Dementia-free Death Before Age 90 Stratified by APOE ε4 Carrier Status

| | APOE ε4 Non-carriers | | | APOE ε4 carriers | | | |
|---|---|---|---|---|---|---|---|
| | | Without Cognitive Impairment (N = 447) | With Cognitive Impairment (N = 356) | | Without Cognitive Impairment (N = 73) | With Cognitive Impairment (N = 145) | |
| Biomarker | SD | OR (95% CI) | OR (95% CI) | SD | OR (95% CI) | OR (95 CI) | P-Interaction |
| p-tau181 | 0.51 | 0.87 (0.71, 1.07) | 1.06 (0.81, 1.39) | 0.53 | 0.77 (0.52, 1.13) | 0.88 (0.55, 1.43) | 0.54 |
| p-tau217 | 0.72 | 0.63 (0.50, 0.80) | 0.93 (0.69, 1.26) | 0.84 | 0.37 (0.23, 0.60) | 0.80 (0.48, 1.35) | 0.13 |
| NfL | 0.56 | 0.71 (0.56, 0.90) | 1.00 (0.73, 1.35) | 0.56 | 0.93 (0.60, 1.42) | 0.66 (0.39, 1.13) | 0.13 |
| GFAP | 0.62 | 0.81 (0.64, 1.02) | 1.20 (0.89, 1.62) | 0.60 | 0.48 (0.30, 0.77) | 0.73 (0.43, 1.25) | 0.068 |

Abbreviations: ADRD, Alzheimer's disease and related dementias; SD, standard deviation; OR, odds ratio; CI, confidence intervals; p-tau181, tau phosphorylated at threonine 181; p-tau217, tau phosphorylated at threonine 217, GFAP, glial fibrillary acidic protein.

f. Participants with missing cognitively healthy longevity outcome or APOE ε4 variables were excluded.
g. The reference group was dementia-free death before age 90 (Non-carrier: N = 249; carrier: N = 119).
h. Model adjusted for the following baseline covariates: age, hormone therapy arm, education, smoking, physical activity, BMI, diabetes, cardiovascular disease, non-melanoma cancer, estimated glomerular filtration rate, total cholesterol, high-density lipoprotein cholesterol, and, hypertension.
i. P-values for interactions were derived from likelihood ratio tests comparing nested models with and without the interaction term between plasma biomarker and APOE ε4.
j. Results represent 1-SD increase in the log2 of plasma biomarkers. All models incorporated the product of inverse propensity score weights and sampling weights.

Table S8. Associations of Plasma ADRD Biomarkers with Odds of Surviving to Age 90 With or Without Cognitive Impairment, Versus Dementia-free Death Before Age 90 Stratified by Randomization to Estrogen Alone vs Placebo

| | | Estrogen alone | | Placebo | | |
|---|---|---|---|---|---|---|
| | | Without Cognitive Impairment (N = 139) | With Cognitive Impairment (N = 167) | Without Cognitive Impairment (N = 158) | With Cognitive Impairment (N = 143) | |
| Biomarker | SD | OR (95% CI) | OR (95% CI) | OR (95% CI) | OR (95% CI) | P-Interaction |
| p-tau181 | 0.55 | 0.74 (0.46, 1.17) | 1.04 (0.74, 1.46) | 0.78 (0.59, 1.04) | 1.04 (0.90, 1.21) | 0.96 |
| p-tau217 | 0.80 | 0.61 (0.38, 0.98) | 1.26 (0.88, 1.80) | 0.55 (0.41, 0.74) | 0.94 (0.81, 1.09) | 0.62 |
| NfL | 0.60 | 0.76 (0.48, 1.20) | 0.96 (0.68, 1.34) | 0.89 (0.67, 1.18) | 0.86 (0.74, 0.99) | 0.54 |
| GFAP | 0.64 | 0.80 (0.50, 1.26) | 1.18 (0.84, 1.65) | 0.88 (0.67, 1.17) | 0.90 (0.78, 1.04) | 0.35 |

Abbreviations: ADRD, Alzheimer's disease and related dementias; SD, standard deviation; OR, odds ratio; CI, confidence intervals; p-tau181, tau phosphorylated at threonine 181; p-tau217, tau phosphorylated at threonine 217, GFAP, glial fibrillary acidic protein.

a. Participants with missing cognitively healthy longevity outcome, race, or ethnicity variables were excluded.
b. The reference group was dementia-free death before age 90 (Intervention: N = 176; Placebo: N = 181).
c. Model adjusted for the following baseline covariates: age, race, ethnicity, education, smoking, physical activity, BMI, diabetes, cardiovascular disease, non-melanoma cancer, estimated glomerular filtration rate, total cholesterol, high-density lipoprotein cholesterol, and hypertension.
d. P-values for interactions were derived from likelihood ratio tests comparing nested models with and without the interaction term between plasma biomarker and intervention arm.
e. Results represent 1-SD increase in the log2 of plasma biomarkers. All models incorporated the product of inverse propensity score weights and sampling weights.

Table S9. Associations of Plasma ADRD Biomarkers with Odds of Surviving to Age 90 With or Without Cognitive Impairment, Versus Dementia-free Death Before Age 90 Stratified by Randomization to Estrogen plus Progestin versus Placebo

| | | Estrogen plus progestin | | Placebo | | |
|---|---|---|---|---|---|---|
| | | Without Cognitive Impairment (N = 243) | With Cognitive Impairment (N = 230) | Without Cognitive Impairment (N = 261) | With Cognitive Impairment (N = 232) | |
| Biomarker | SD | OR (95% CI) | OR (95% CI) | OR (95% CI) | OR (95% CI) | P-Interaction |
| p-tau181 | 0.54 | 0.82 (0.56, 1.19) | 1.24 (0.92, 1.67) | 0.85 (0.65, 1.10) | 0.98 (0.84, 1.14) | 0.40 |
| p-tau217 | 0.77 | 0.59 (0.39, 0.87) | 0.99 (0.73, 1.34) | 0.55 (0.42, 0.71) | 0.92 (0.79, 1.08) | 0.93 |
| NfL | 0.57 | 0.64 (0.44, 0.93) | 1.00 (0.74, 1.34) | 0.62 (0.48, 0.81) | 0.65 (0.56, 0.76) | 0.12 |
| GFAP | 0.63 | 0.79 (0.54, 1.15) | 1.24 (0.92, 1.67) | 0.69 (0.53, 0.89) | 0.90 (0.78, 1.05) | 0.40 |

Abbreviations: ADRD, Alzheimer's disease and related dementias; SD, standard deviation; OR, odds ratio; CI, confidence intervals; p-tau181, tau phosphorylated at threonine 181; p-tau217, tau phosphorylated at threonine 217, GFAP, glial fibrillary acidic protein.

a. Participants with missing cognitively healthy longevity outcome, race, or ethnicity variables were excluded.
b. The reference group was dementia-free death before age 90 (Intervention: N = 210; Placebo: N = 194).
c. Model adjusted for the following baseline covariates: age, race, ethnicity, education, smoking, physical activity, BMI, diabetes, cardiovascular disease, non-melanoma cancer, estimated glomerular filtration rate, total cholesterol, high-density lipoprotein cholesterol, and hypertension.
d. P-values for interactions were derived from likelihood ratio tests comparing nested models with and without the interaction term between plasma biomarker and intervention arm.
e. Results represent 1-SD increase in the log2 of plasma biomarkers. All models incorporated the product of inverse propensity score weights and sampling weights.

Table S10. Associations of Plasma ADRD Biomarkers with Odds of Surviving to Age 90 With or Without Cognitive Impairment, Versus Dementia-free Death Before Age 90 Stratified After Removing Outliers (5-SD above the Mean)

| | | Without cognitive Impairment | | With cognitive impairment | |
|---|---|---|---|---|---|
| Biomarker | SD | OR (95% CI) | P-value | OR (95% CI) | P-value |
| p-tau181 | 0.54 | 0.81 (0.71, 0.93) | 0.0023 | 1.05 (0.88, 1.24) | 0.61 |
| p-tau217 | 0.78 | 0.58 (0.50, 0.68) | <0.0001 | 1.01 (0.84, 1.22) | 0.90 |
| NfL | 0.58 | 0.71 (0.61, 0.82) | <0.0001 | 0.84 (0.69, 1.02) | 0.079 |
| GFAP | 0.64 | 0.78 (0.68, 0.91) | 0.0009 | 1.04 (0.86, 1.25) | 0.68 |

Abbreviations: ADRD, Alzheimer's disease and related dementias; SD; standard deviation; OR, odds ratio; CI, confidence intervals; p-tau181, tau phosphorylated at threonine 181; p-tau217, tau phosphorylated at threonine 217; NfL, neurofilament light; GFAP, glial fibrillary acidic protein.

a. Participants with missing cognitively healthy longevity outcome, race, ethnicity variables, or if their plasma biomarker levels were more than 5-SD above the mean were excluded.
b. The reference group was dementia-free death before age 90.
c. Model adjusted for the following baseline covariates: age, race, ethnicity, hormone therapy arm, education, smoking, physical activity, BMI, diabetes, cardiovascular disease, non-melanoma cancer, estimated glomerular filtration rate, total cholesterol, high-density lipoprotein cholesterol, and hypertension.
d. Results represent 1-SD increase in the log2 of plasma biomarkers. All models incorporated the product of inverse propensity score weights and sampling weights.

Table S11. Associations of Plasma ADRD Biomarkers with Odds of Surviving to Age 90 With or Without Cognitive Impairment Versus Death Before Age 90, Including Dementia Deaths.

| | | Without cognitive Impairment | | With cognitive impairment | |
|---|---|---|---|---|---|
| Biomarker | SD | OR (95% CI) | P-value | OR (95% CI) | P-value |
| p-tau181 | 0.55 | 0.78 (0.68, 0.88) | 0.0001 | 1.02 (0.86, 1.21) | 0.82 |
| p-tau217 | 0.82 | 0.52 (0.44, 0.60) | <0.0001 | 0.90 (0.75, 1.09) | 0.29 |
| NfL | 0.58 | 0.70 (0.61, 0.81) | <0.0001 | 0.84 (0.69, 1.01) | 0.061 |
| GFAP | 0.63 | 0.74 (0.64, 0.85) | <0.0001 | 0.98 (0.82, 1.17) | 0.82 |

Abbreviations: ADRD, Alzheimer's disease and related dementias; SD, standard deviation; OR, odds ratio; CI, confidence intervals; p-tau181, tau phosphorylated at threonine 181; p-tau217, tau phosphorylated at threonine 217, GFAP, glial fibrillary acidic protein.

a. Participants with missing cognitively healthy longevity outcome, race, ethnicity variables were excluded.
b. The reference group was death before age 90 (including both dementia and dementia-free deaths)
c. Model adjusted for the following baseline covariates: age, race, ethnicity, hormone therapy arm, education, smoking, physical activity, BMI, diabetes, cardiovascular disease, non-melanoma cancer, estimated glomerular filtration rate, total cholesterol, high-density lipoprotein cholesterol, and hypertension.
d. Results represent 1-SD increase in the log2 of plasma biomarkers. All models incorporated the product of inverse propensity score weights and sampling weights.
